# Small volumes, deep insights: longitudinal plasma EV multi-omics in very preterm infants

**DOI:** 10.64898/2026.02.04.26345553

**Authors:** Nicole Grinovero, Francesca Antonini, Martina Bartolucci, Lucilla Rossi, Gianvittorio Luria, Maurizio Bruschi, Sonia Spinelli, Gino Tripodi, Chiara Andreato, Francesco Vinci, Luca Antonio Ramenghi, Chiara Lavarello, Andrea Petretto

**Affiliations:** Core Facility for Omics Sciences, IRCCS Istituto Giannina Gaslini, Genoa, Italy; Unit of Nephrology, Dialysis, and Transplantation and Laboratory of Molecular Nephrology, IRCCS Istituto Giannina Gaslini, Genoa, Italy; Department of Experimental Medicine (DIMES), University of Genoa, Italy; Immunohematology and Transfusion Medicine Unit, IRCCS Istituto Giannina Gaslini, Genoa, Italy; Department of Neuroscience, Rehabilitation, Ophthalmology, Genetics, Maternal and Child Health, University of Genoa, Italy; Neonatal Intensive Care Unit, IRCCS Istituto Giannina Gaslini, Genoa, Italy

**Author notes:** Correspondence to: Chiara Lavarello. These authors contributed equally to this work (Nicole Grinovero, Francesca Antonini). These authors jointly supervised this work (Luca Antonio Ramenghi, Chiara Lavarello, Andrea Petretto).

**Keywords:** Extracellular vesicles, Multi-omics, Preterm birth, Liquid biopsy, Brain injury

## Abstract

Very preterm birth disrupts critical fetal developmental programs, yet the systemic molecular trajectories driving extrauterine adaptation remain poorly defined. Although extracellular vesicles (EVs) represent informative systemic compartments, comprehensive multi-omics is constrained by the small plasma volumes safely obtainable from neonates. Here, we adapted a magnetic bead-based framework (Mag-Net) to enable parallel EV proteomics and lipidomics from the same EV-enriched preparation using 10 µL of plasma. Across 74 longitudinal samples collected from birth to term-equivalent age, we quantified 1,528 EV-associated proteins and 421 lipid species. The EV proteome shifted from early translation and metabolic programs toward progressive immune competence, while the lipidome underwent selective structural remodeling enriched in triacylglycerols and ether-linked phosphatidylcholines. Cross-omics integration identified coordinated protein-lipid modules associated with clinical phenotypes, including brain injury. This study demonstrates that parallel EV proteomic-lipidomic profiling from microliter plasma volumes is feasible and captures coordinated developmental and clinically relevant programs in very preterm infants.

## Introduction

Preterm birth affects approximately 13-15 million infants each year worldwide and remains a leading cause of neonatal mortality and long-term neurodevelopmental impairment^1^. Infants born very preterm (<32 weeks of gestation) undergo extrauterine adaptation during the third trimester, a critical window marked by the rapid and coordinated maturation of neural, immune and metabolic systems^2,3^. This transition interrupts tightly regulated fetal programs, forcing maturation to unfold under fundamentally different biological conditions shaped by altered oxygen exposure, inflammation, and modified nutritional inputs. As a result, postnatal development in very preterm infants does not simply represent an accelerated version of normal maturation, but unfolds under different biological constraints^4,5^.

During the first weeks of life, intrinsic maturation occurs alongside clinical exposures such as respiratory support, infection and hemodynamic instability. Because developmental timing and stress responses are tightly intertwined, cross-sectional comparisons provide limited insight into the molecular organization of early adaptation. In addition, inter-individual variability is substantial in very preterm cohorts, reflecting differences in gestational age and clinical course. Longitudinal designs, in which each infant serves as their own reference, are therefore essential to distinguish structured developmental trajectories from reactive changes linked to clinical events^6^.

Extracellular vesicles (EVs) provide a biologically relevant analytically tractable compartment for this purpose. EVs are membrane-bound particles released by most cell types and mediate intercellular communication through the transfer of proteins, lipids and nucleic acids^7,8^. As circulating, cell-derived structures, they integrate signals across tissues and can be accessed through minimally invasive plasma sampling, making them well suited for longitudinal studies in neonatal populations^9,10^. Importantly, EVs are structured molecular entities. Their lipid bilayer is selectively assembled and enriched in specific phospholipids and sphingolipids that influence vesicle curvature, stability and fusion, while membrane-associated and luminal proteins regulate trafficking and signaling^11^. Because membrane lipids and proteins are functionally linked, parallel analysis of EV proteomes and lipidomes enables an integrated view of both structural and signaling aspects of systemic adaptation.

Despite this potential, EV analysis in plasma poses well-recognized technical challenges. EVs represent only a small fraction of circulating particles and are outnumbered by lipoproteins and abundant soluble proteins. Overlap in size and density complicates separation, and incomplete resolution is particularly problematic for lipidomics, where co-isolated lipoprotein lipids can dominate the measured signal^12,13^. Benchmarking studies have shown that isolation strategies involve trade-offs between purity and yield: precipitation-based methods often provide higher recovery but increased contamination, whereas density-gradient or size-exclusion approaches improve purity but reduce yield and require larger input volumes^12^.

These limitations are amplified in neonatal research, where available plasma volumes are inherently limited and repeated sampling must remain minimally invasive to allow longitudinal follow-up. Many EV isolation workflows require hundreds of microliters to milliliters of plasma and multiple processing steps, making them poorly suited for fragile populations. In addition, most EV multi-omics studies process proteomics and lipidomics separately, dividing enriched material into aliquots. When starting volumes are small, this reduces analytical depth and disrupts the natural coupling between membrane composition and protein cargo. Although parallel EV proteomics and lipidomics have been demonstrated in adult plasma using larger-volume, density-gradient-based workflows^13^, integrated proteomic-lipidomic profiling from microliter-scale plasma inputs in a longitudinal neonatal setting has not been established.

To address part of the isolation challenge, a strong anion exchange (SAX)-based enrichment strategy (Mag-Net) was recently developed for plasma proteomics^14^. By exploiting the net negative surface charge of EV membranes, Mag-Net enables selective enrichment of vesicle-associated material from complex plasma matrices while reducing interference from highly abundant soluble proteins. This workflow has been validated for deep plasma EV proteome profiling and provides an automated, scalable platform compatible with clinical samples^14^.

In this study, we build on the Mag-Net framework and extend it to enable integrated proteomic and lipidomic analysis from plasma micro-volumes. We evaluate and refine the workflow using 5 and 10 µL plasma inputs, volumes compatible with longitudinal sampling in very preterm infants, and optimize the downstream extraction strategy to enable monophasic recovery of proteins and lipids from the same EV preparation. This approach preserves cross-layer molecular relationships without splitting the input material and demonstrates the feasibility of parallel EV multi-omics from microliter-scale plasma volumes.

After technical validation and benchmarking against conserved circulating EV signatures^13^, we apply this optimized workflow longitudinally in a clinically characterized cohort of very preterm infants from birth to term-equivalent age. Using linear mixed-effects modeling and cross-omics network integration, we characterize coordinated EV molecular trajectories during early extrauterine adaptation and examine their associations with clinical outcomes, with particular attention to brain injury.

## Results

### Optimization of an integrated EV proteomic and lipidomic workflow from plasma micro-volumes

Integrated clinical multi-omics in fragile populations, such as neonates or patients with rare diseases, is often constrained by minimal sample availability. To enable simultaneous protein and lipid profiling under these micro-volume conditions, we adapted the previously established Mag-Net workflow^14^ – originally validated for plasma proteomics – to support parallel analysis from as little as 5-10 µL of plasma. While preserving the upstream strong anion exchange (SAX)-based EV enrichment step, optimization focused on the downstream extraction strategy to ensure compatibility with both analytes. We compared three monophasic extraction chemistries: methanol (MeOH), isopropanol (IPA), and a 1-butanol/acetonitrile/water mixture (3:1:1, v/v), based on the Bead-enabled Accelerated Monophasic Multi-omics (BAMM) approach^15^. In all conditions, solvent-mediated membrane disruption was coupled with on-bead protein aggregation capture (PAC), allowing coordinated recovery of lipids and proteins from the same EV-enriched preparation.

Proteomic depth remained stable across extraction strategies and input volumes (**Fig. 1a**). At 10 µL of plasma input, we identified 866-992 protein groups, with only minor losses in coverage when scaling down to 5 µL. In contrast, direct digestion of whole plasma (“neat”) yielded markedly lower depth (308 protein groups), confirming that SAX-based enrichment significantly increases the detectable complexity of the EV-associated proteome. Quantitative reproducibility was consistently high, with most proteins exhibiting a coefficient of variation (CV) < 20% (**Fig. 1b**). Furthermore, concordance with the standard Mag-Net implementation remained strong (Pearson *r* = 0.89-0.95; **Fig. 1c**), and the preservation of the global EV-associated proteomic landscape was further supported by correlation structure and protein overlap analysis (**Fig. S1**).

**Fig. 1:**
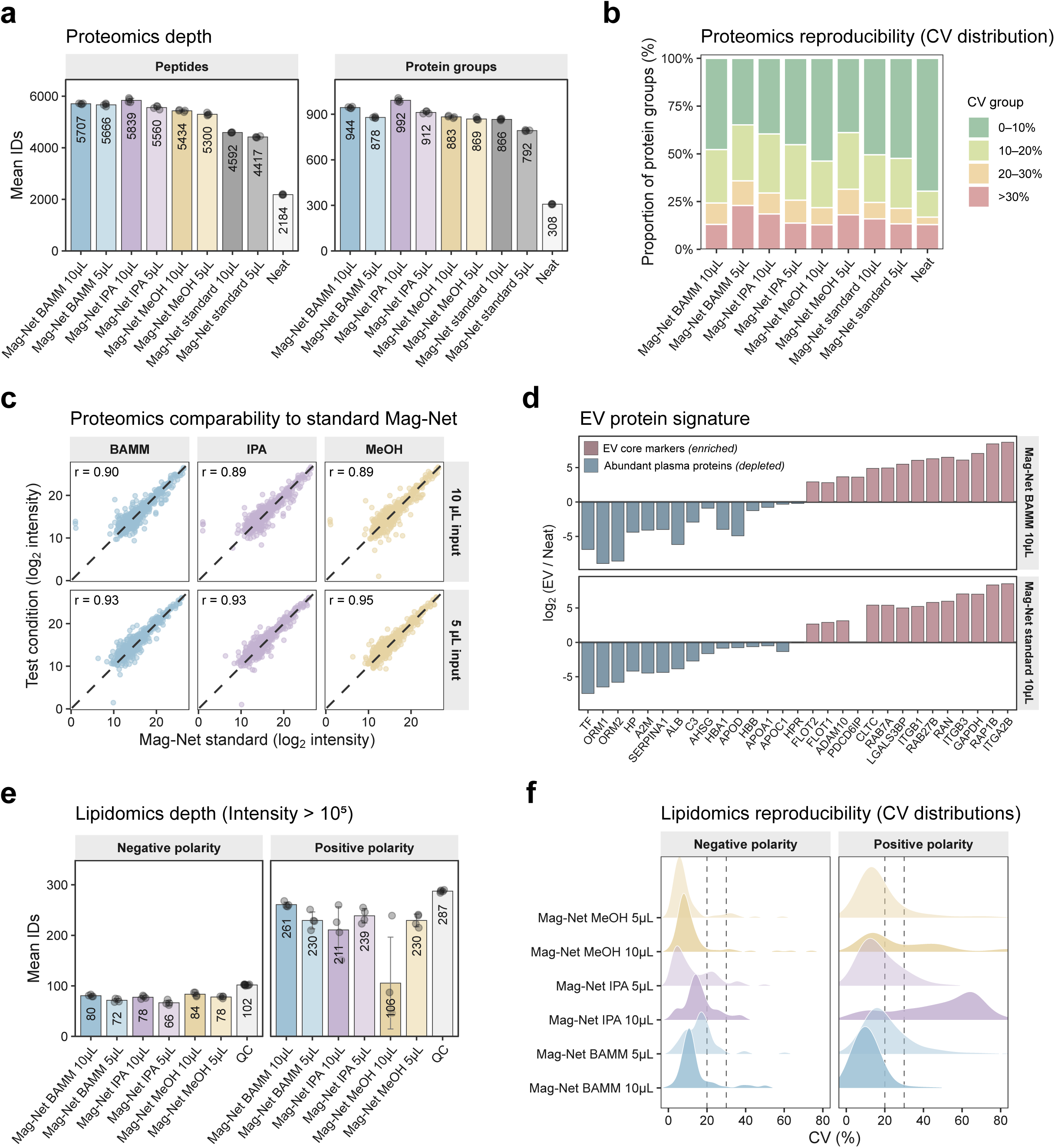
Technical validation of the EV-based multi-omics workflow using plasma micro-volumes. (a) Mean peptide and protein group identifications obtained from 5 µL and 10 µL plasma inputs following EV enrichment and monophasic solvent extraction with 1-butanol/acetonitrile/water (3:1:1, v/v; BAMM), isopropanol (IPA), or methanol (MeOH), compared to the standard Mag-Net protocol. Dots represent technical replicates; bars indicate mean values. “Neat” denotes direct plasma digestion without EV enrichment. (b) Distribution of coefficients of variation (CV) for quantified protein groups across conditions. Bars indicate the proportion of proteins within predefined CV ranges. (c) Scatter plots of log₂-transformed protein intensities comparing each extraction condition with the corresponding standard Mag-Net reference at matched plasma input. Pearson correlation coefficients (r) are shown; dashed lines indicate the identity line. (d) Log₂ fold change (EV/Neat) is shown for representative enriched EV core markers and depleted abundant plasma proteins under BAMM 10 µL and Mag-Net standard 10 µL conditions. (e) Lipidomic depth after application of an intensity threshold (>1×10□). Mean numbers of detected lipid species in negative and positive ionization modes across extraction strategies and plasma input volumes. Dots represent technical replicates; bars indicate mean values. QC denotes pooled quality control samples. (f) Distribution of CV (%) for quantified lipid species in negative and positive polarity. Dashed vertical lines indicate 20% and 30% CV thresholds.

Conversely, lipidomic performance was more sensitive to the choice of extraction solvent. Across conditions, we detected up to 308 lipid species in positive mode and 108 in negative mode (**Fig. S2**). While total feature counts were broadly comparable, restricting the analysis to quantitatively robust signals (intensity > 1×10□) revealed solvent-dependent differences (**Fig. 1e**; **Fig. S2**). BAMM extraction consistently yielded the highest lipid coverage in positive mode and the lowest CV values (**Fig. 1f**), reflecting superior quantitative reproducibility. Notably, lipid class distributions remained stable across solvents (**Fig. S2**), indicating that extraction chemistry primarily influenced detection depth rather than inducing systematic compositional bias.

To verify that these molecular profiles reflect selective EV enrichment rather than plasma carryover, we benchmarked our dataset against a conserved circulating EV atlas defined by Rai et al. using density-gradient fractionation^13^. Of the 182 highly conserved EV proteins reported in that study, 168 (92%) were identified in our dataset. These markers showed coherent enrichment in Mag-Net preparations relative to direct plasma digestion, while abundant non-EV plasma proteins were strongly depleted (**Fig. 1d**). At the lipid level, 9 of the 52 conserved EV lipid features described in the reference atlas were detected; although the absolute overlap was lower than for proteins, these features were consistently detected across all Mag-Net conditions, with BAMM 10 µL displaying the most robust profile (**Fig. S2**). Collectively, these data demonstrate that the adapted Mag-Net-based workflow recapitulates a conserved EV molecular signature, with BAMM extraction at 10 µL input providing the optimal balance of lipidomic robustness and proteomic integrity.

### Longitudinal evolution of the plasma EV proteome and lipidome across very preterm development

With the optimized EV multi-omics workflow established, we investigated how circulating EV molecular programs evolve during early extrauterine adaptation. We applied the method to a longitudinal cohort of 16 very preterm infants (<32 weeks gestational age) followed from birth to term-equivalent age (TEA; 40 weeks postmenstrual age). TEA serves as a critical clinical reference point, representing the stage at which a preterm infant reaches the developmental maturity of a term-born newborn.

Plasma samples were collected at five predefined timepoints: birth (T0), 48-72 hours (T1), 7 days (T2), 33 weeks postmenstrual age (T3), and TEA (T4). While all 16 infants contributed samples for the T0-T3 interval, 10 were followed up to the final TEA milestone. The longitudinal design and analytical workflow are summarized in **Fig. 2a**, and clinical characteristics are reported in **Table 1**. Parallel analysis of proteins and lipids from each sample enabled within-subject tracking of coordinated multi-omic trajectories over time. Based on the superior coverage and quantitative robustness observed during our technical benchmarking, longitudinal lipidomic profiling was prioritized in positive ionization mode to ensure maximum analytical depth across the cohort.

**Fig. 2:**
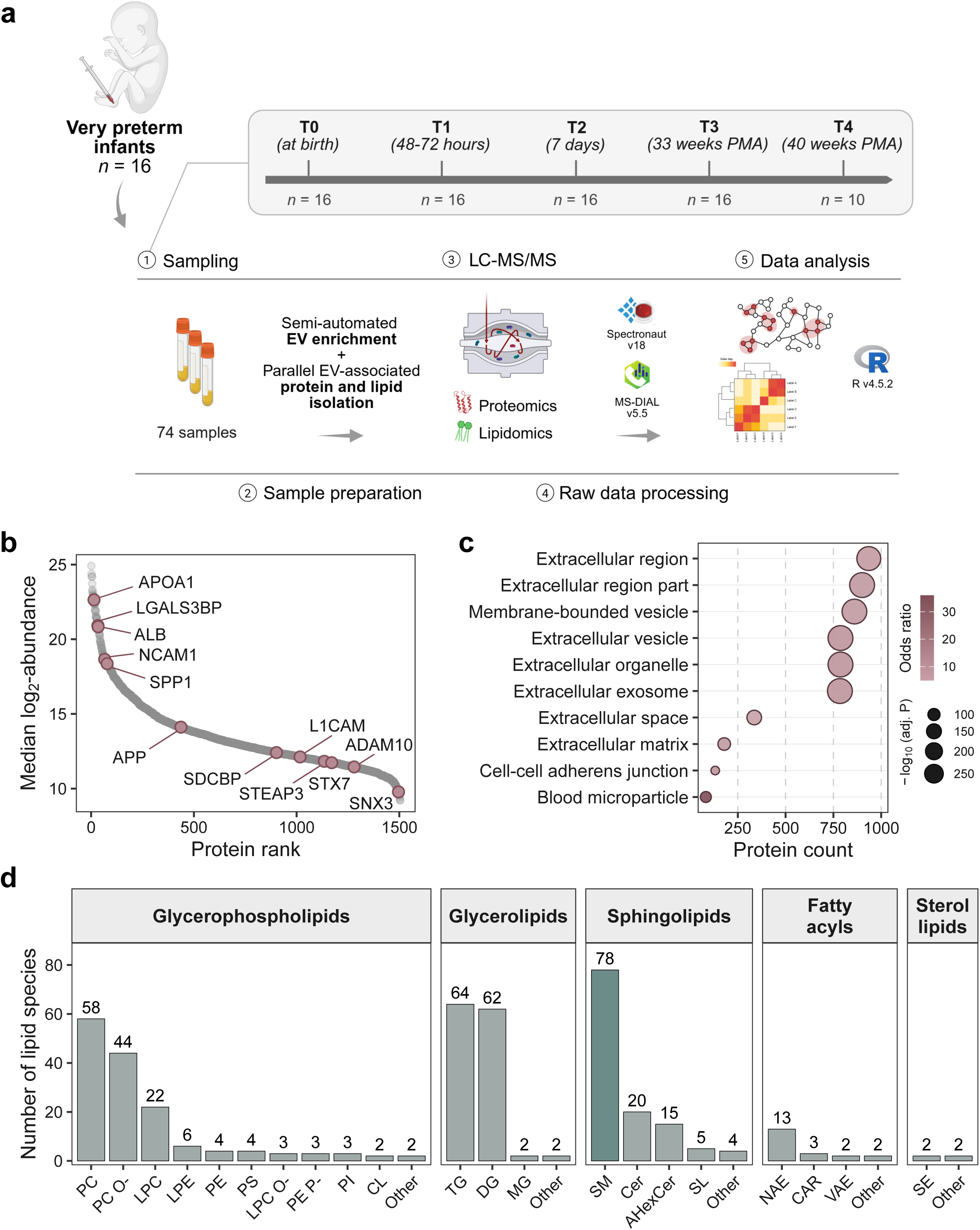
Longitudinal study design and plasma EV multi-omics profiling in very preterm infants. (a) Overview of the longitudinal cohort, sampling timeline, and analytical workflow. Plasma samples were collected from very preterm infants born at <32 weeks of gestational age (n = 16) at five timepoints spanning birth to term-equivalent age: birth (T0), 48-72 hours (T1), 7 days (T2), 33 weeks postmenstrual age (T3), and 40 weeks postmenstrual age (T4, n = 10). EVs were enriched from plasma, followed by parallel isolation of EV-associated proteins and lipids from the same sample. Proteomic and lipidomic profiles were generated by LC-MS/MS and processed through standardized pipelines. (b) Dynamic range of the plasma EV proteome shown as a rank-abundance plot of median log₂ protein abundance across all samples, highlighting representative EV-associated proteins. (c) Subcellular localization enrichment analysis of the EV proteome based on the Jensen COMPARTMENTS database. Terms are ordered by associated protein count along the x axis. Dot size reflects statistical significance (−log_10_ FDR), while color intensity indicates odds ratio. (d) Distribution of identified lipid species across major lipid categories, including glycerophospholipids, glycerolipids, sphingolipids, fatty acyls, and sterol lipids.

**Table 1:**
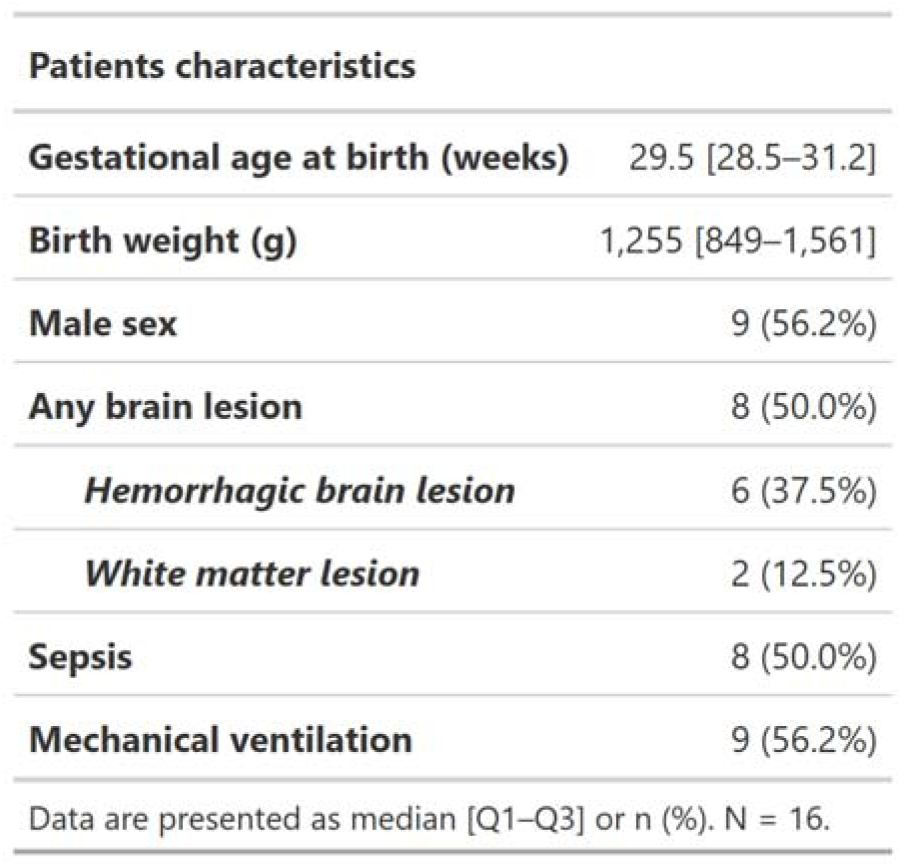
Baseline demographic and clinical characteristics of the study cohort (N = 16). Continuous variables are reported as median [Q1–Q3] and categorical variables as n (%). Neurological outcomes are presented hierarchically, with overall brain lesion status reported as the primary outcome and lesion subtypes described separately.

After quality filtering, we retained 1,528 EV-associated protein groups and 421 unique lipid species for longitudinal analysis. Post-filter quantification depth per sample remained stable across timepoints in both omics layers (**Fig. S3**), supporting consistent analytical coverage throughout the longitudinal series. The EV-associated proteome spanned a wide dynamic range (**Fig. 2b**) and showed significant over-representation of extracellular and vesicle-related compartments, consistent with its EV origin (**Fig. 2c**). In parallel, the lipidomic profile revealed a membrane-enriched composition dominated by glycerophospholipids and sphingolipids, with sphingomyelins as the predominant lipid class (**Fig. 2d**).

To characterize the systematic remodeling of proteomic and lipidomic profiles, we employed a linear mixed-effects modeling (LMM) framework. This approach accounted for the longitudinal design by incorporating infant-specific random intercepts to model within-subject correlations over time. Sampling timepoints (T0-T4) were treated as fixed effects to capture developmental trends, while gestational age at birth was included as a covariate, as it represents a major source of biological heterogeneity in very preterm cohorts and strongly influences baseline molecular profiles. For each molecular feature, we tested for a global effect of time and quantified the primary developmental shift by contrasting TEA (T4) against birth (T0), using the Benjamini-Hochberg (BH) procedure to control for multiple testing.

### Extensive postnatal remodeling of the circulating EV proteome

Applying the LMM framework revealed a profound temporal transformation of the plasma EV proteome following very preterm birth. Overall, 57% of the quantified proteins (875 of 1,528) exhibited a significant global effect of time (FDR < 0.05), indicating that the majority of EV cargo is actively and dynamically regulated during the first weeks of extrauterine life.

Unsupervised hierarchical clustering of these time-regulated proteins resolved four distinct clusters that follow two primary, opposing trajectories: proteins that progressively decreased from birth toward maturation (clusters 1A and 1B) and proteins that steadily increased over time (clusters 2A and 2B) (**Fig. 3a,b**). The presence of these highly coordinated trajectories suggests that longitudinal EV proteomic shifts are coherent and directional, rather than reflecting unspecific physiological variability.

**Fig. 3:**
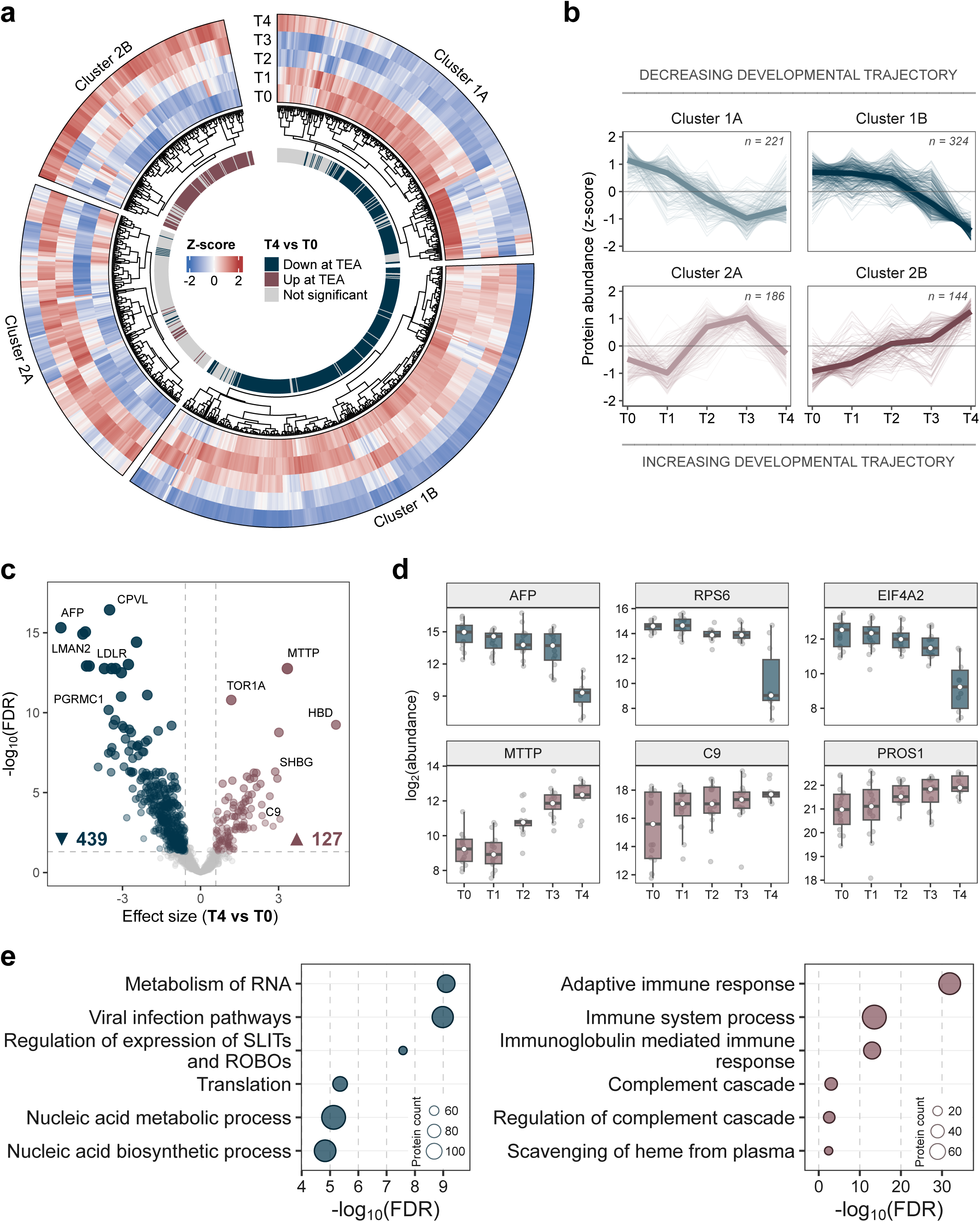
Coordinated longitudinal remodeling of the plasma EV proteome from birth to term-equivalent age. (a) Circular heatmap illustrating the 875 EV proteins with a significant temporal profile (LMM; FDR < 0.05). Data are shown as row z-scores of median protein abundances across five postnatal timepoints, from birth (T0) to term-equivalent age (TEA, T4). Hierarchical clustering resolved four primary modules; the outer ring indicates the direction of the net developmental shift (T4 vs T0). (b) Cluster-specific temporal trajectories. Profile plots show z-scored abundances for individual proteins (thin lines) and cluster medians (bold lines), highlighting coordinated decreasing (clusters 1A, 1B) and increasing (clusters 2A, 2B) proteomic programs (n indicates protein count per cluster). (c) Volcano plot displaying the differential protein abundance between TEA (T4) and birth (T0) derived from the LMM. Statistical significance (−log10 FDR) is plotted against the effect size (β coefficient). Significant features (FDR < 0.05 and |β| > 0.58) are highlighted, with counts of downregulated (blue, 439) and upregulated (red, 127) proteins indicated. (d) Longitudinal profiles of representative EV-associated proteins. Boxes represent median and interquartile range; whiskers indicate data range. (e) Functional enrichment analysis of significantly downregulated (left) or upregulated proteins (right) at TEA. Top enriched terms from Gene Ontology (Biological Process) and Reactome databases are ranked by statistical significance (−log10 FDR). Dot size reflects the number of proteins associated with each term.

To quantify the magnitude of this transition, we focused on the net developmental shift between birth and TEA. We identified 439 proteins significantly downregulated and 127 proteins significantly upregulated at TEA (FDR < 0.05 and |β| > 0.58, corresponding to approximately ≥1.5-fold change; **Fig. 3c**). Proteins that decreased toward TEA were central to RNA metabolism, translation, and early growth-related processes (**Fig. 3e**). This “early-life” signature included core components of the translational machinery, such as RPS6 and EIF4A2, alongside the classic fetal marker alpha-fetoprotein (AFP), all of which showed steep declines across the five timepoints (**Fig. 3d**).

In striking contrast, the proteomic profile at TEA was characterized by a mature systemic signature. Proteins increasing toward TEA were significantly enriched for immune-related functions, specifically the complement cascade, immunoglobulin-mediated responses, and adaptive immunity (**Fig. 3e**). Representative markers of this maturation – including complement component C9, the anticoagulant protein S (PROS1), and the lipid transporter MTTP – exhibited linear increases across postnatal development (**Fig. 3d**). Collectively, these findings highlight a coordinated functional transition of the EV proteome from a translation-heavy growth program at birth toward a robust immune-competent profile at term-equivalent age.

### Selective structural and metabolic remodeling of the EV lipidome

We next evaluated the longitudinal evolution of the plasma EV lipidome using the same LMM framework. While lipidomic remodeling involved a more targeted subset of features compared to the proteome – with 100 of 421 quantified species showing a significant global effect of time (FDR < 0.05) – the changes were highly structured and coordinated (**Fig. 4a**). Unsupervised hierarchical clustering resolved these regulated lipids into four distinct developmental trajectories, each comprising species from multiple chemical classes (**Fig. 4b**). This modular organization indicates that EV lipidomic shifts are governed by coordinated metabolic programs rather than uniform, class-wide abundance changes.

**Fig. 4:**
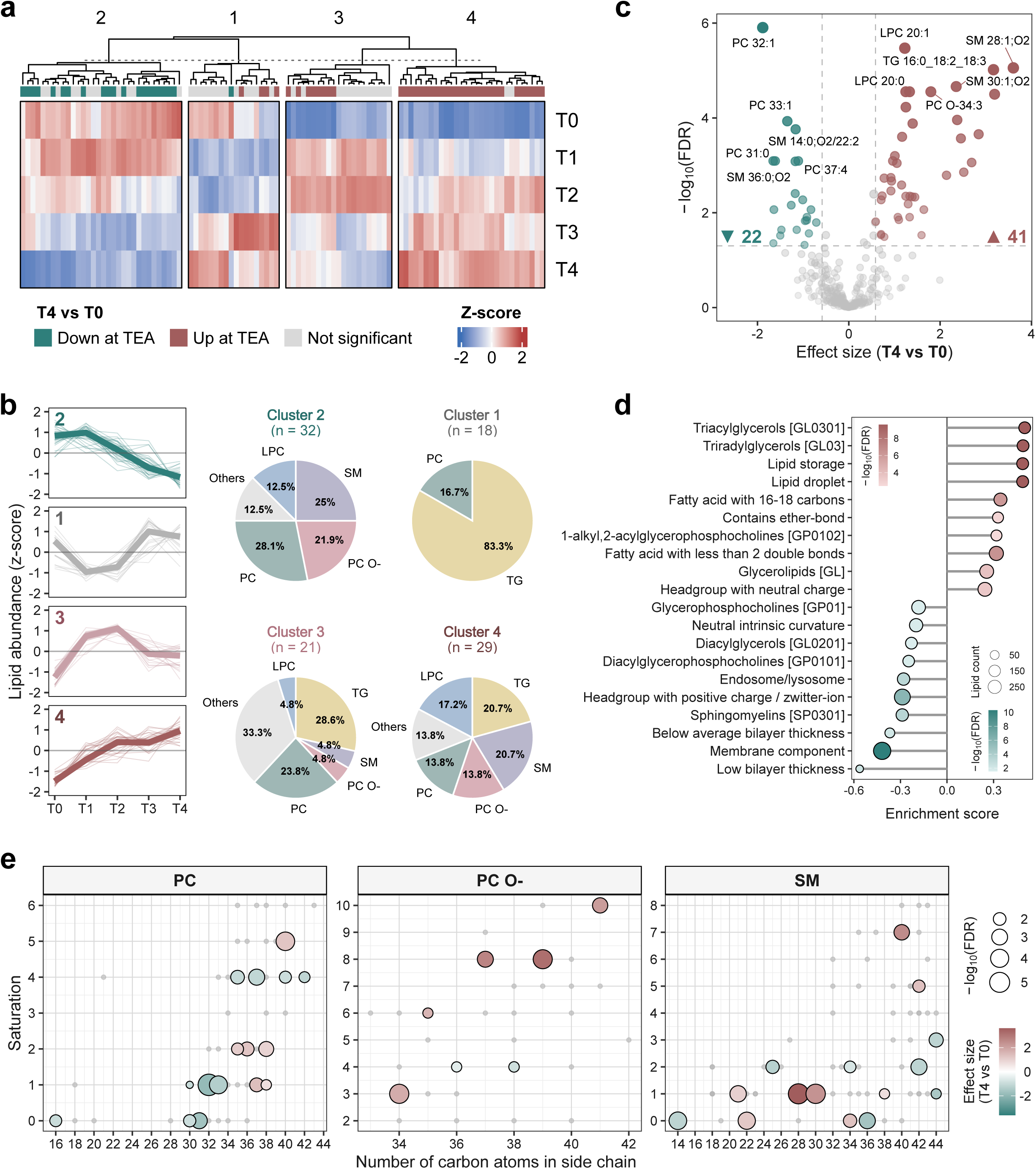
Coordinated structural and metabolic maturation of the plasma EV lipidome. (a) Heatmap of the 100 lipid species showing a significant temporal profile in the longitudinal linear mixed-effects model (LMM; FDR < 0.05). Data are represented as row z-scores of median abundances across five postnatal timepoints (T0-T4). Hierarchical clustering identifies four distinct longitudinal trajectories; the right-side annotation indicates the direction of change at term-equivalent age (TEA, T4) relative to birth (T0). (b) Cluster-specific temporal trajectories and compositional analysis. Profile plots (left) show z-scored abundances for individual lipids (thin lines) and cluster medians (bold lines). The distribution of lipid classes within each cluster is shown as pie charts (right). (c) Volcano plot displaying differential lipid abundance between TEA (T4) and birth (T0). Statistical significance (−log10 FDR) is plotted against the effect size (β coefficient). Significant features (FDR < 0.05, |β| > 0.58) are highlighted, with counts of upregulated (red, 41) and downregulated (green, 22) lipids indicated. (d) Lipid ontology (LION) enrichment analysis based on the β coefficient (T4 vs T0). Enrichment scores are represented as lollipop plots, where dot size reflects the number of annotated lipid species and color intensity indicates statistical significance (−log10 FDR). (e) Structural space distribution of regulated phosphatidylcholines (PC), ether-linked phosphatidylcholines (PC O-), and sphingomyelins (SM). Lipids are mapped according to the number of carbons in the side chain (x-axis) and degree of unsaturation (y-axis). Dot size reflects statistical significance (−log10 FDR) and color encodes effect size (T4 vs T0), highlighting selective structural redistribution across postnatal development.

The developmental transition to TEA was marked by the significant upregulation of 41 lipid species and the downregulation of 22 (FDR < 0.05 and |β| > 0.58, corresponding to approximately ≥1.5-fold change; **Fig. 4c**). Lipids increasing over development were dominated by triacylglycerols (TG), ether-linked phosphatidylcholines (PC O-), and specific sphingomyelins (SM). Among the strongest increases were SM 28:1;O2 and ether-linked species such as PC O-39:8 and PC O-37:8, alongside multiple TG species (**Fig. 4c**). These shifts are consistent with a progressive enrichment of neutral lipid cargo and a selective remodeling of EV membrane architecture.

Conversely, lipids that declined toward TEA were significantly enriched for diacyl PCs, phosphatidylserines (PS), diacylglycerols (DG), and long-chain SMs. Representative examples included PC 32:1 and PC 31:0, which showed the strongest negative effect sizes, as well as long-chain SM such as SM 36:0;O2 and SM 44:1;O2. Notably, lipid regulation was highly structural-specific rather than class-uniform. Specifically, SMs displayed opposite trends depending on chain length, with shorter species increasing and longer species decreasing over development, while PCs had divergent behavior according to linkage type, with ether-linked species preferentially increasing and diacyl species preferentially decreasing toward TEA.

To place these compositional changes into a functional context, we performed lipid ontology (LION) enrichment analysis based on the TEA vs birth β coefficients (log₂ fold-change estimates) (**Fig. 4d**). Lipids enriched at T4 were associated with annotations related to lipid storage and lipid droplet biology. In contrast, lipids reduced over time were preferentially associated with membrane structural properties, including bilayer thickness and intrinsic curvature.

Finally, we mapped the regulated species from major membrane classes (PC, PC O-, and SM) onto carbon number-double bond space (**Fig. 4e**). This analysis revealed that birth- and TEA-enriched species occupy distinct regions of structural space within each class. Importantly, these shifts were class-specific and did not reflect a uniform global trend toward increased chain length or saturation, but rather a selective redistribution among molecular species.

Together, these analyses demonstrate that postnatal development in very preterm infants is characterized by a progressive and highly coordinated remodeling of the plasma EV composition across both omics layers. While proteomic changes are widespread and involve fundamental shifts in pathways related to growth and immune maturation, lipidomic changes are more selective, reflecting a sophisticated redistribution among specific structural species. Overall, these findings establish that plasma EVs serve as high-resolution molecular snapshots of the systemic developmental changes occurring during the critical preterm period, providing a robust platform for tracking extrauterine adaptation.

### Network-based integration reveals coordinated multi-omic EV modules

To characterize the systemic architecture of EV maturation, we integrated the proteomic and lipidomic layers using a cross-omics correlation network approach (**Fig. 5a**). We focused on the dynamically regulated components of EV remodeling by including only the 875 proteins and 100 lipids that exhibited significant temporal trajectories in our longitudinal LMM analysis. Inter-omic coordination was assessed by computing pairwise Spearman correlations (ρ) across paired longitudinal samples. By restricting the network to cross-layer associations and to features with a significant time effect (FDR < 0.05), we intentionally captured coordinated protein–lipid co-variation along the shared developmental trajectory, rather than within-layer structure or correlations driven by absolute abundance. After multiple-testing correction and effect-size filtering (|ρ| ≥ 0.35, FDR < 0.05), the resulting network revealed a bimodal distribution of associations, with clear topological separation between positive and negative correlations (**Fig. 5b**).

**Fig. 5:**
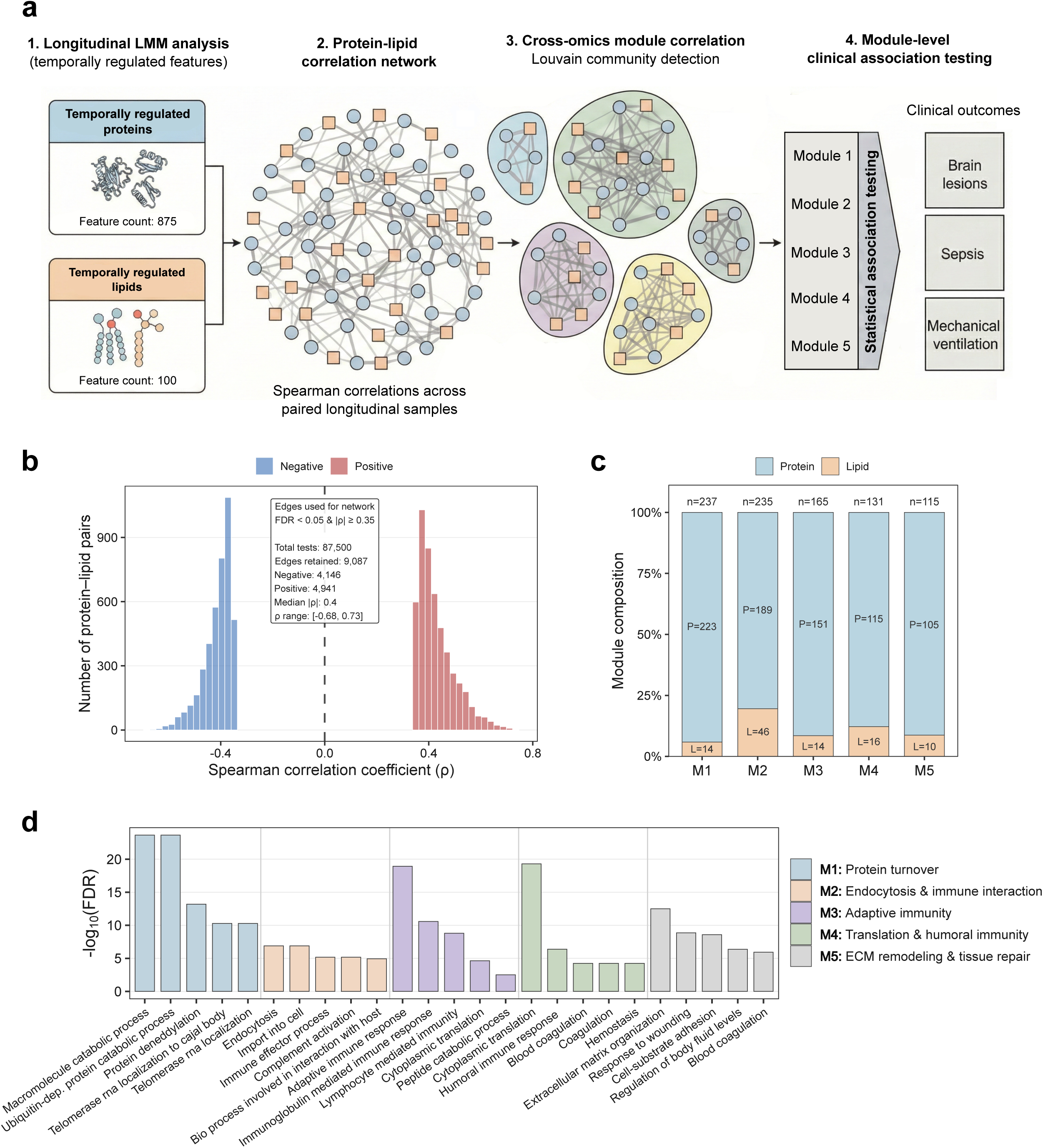
Cross-omics integration of the plasma EV proteome and lipidome. (a) Schematic overview of the cross-omics integration strategy. Temporally regulated EV proteins (n = 875) and lipids (n = 100), identified by longitudinal linear mixed-effects modeling (LMM), were integrated using a protein-lipid correlation network based on Spearman correlations across paired longitudinal samples. Louvain community detection was applied to identify cross-omics modules, which were subsequently tested for associations with clinical outcomes. (b) Distribution of Spearman correlation coefficients (ρ) for all protein-lipid pairs. Only significant edges (FDR < 0.05 and |ρ| ≥ 0.35) were retained for network construction. (c) Composition of the five cross-omics modules (M1-M5) by molecular layer (protein vs lipid). Bars indicate the relative contribution of proteins and lipids within each module, with total node counts reported above each bar. (d) Functional enrichment analysis of the protein components within each module based on the Gene Ontology Biological Process database. Terms are ranked by −log10(FDR) and colored by module assignment.

Using Louvain community detection, we resolved the network into five distinct cross-omic modules (M1-M5), each representing a set of proteins and lipids with coherent longitudinal behavior (**Fig. 5a, c**). While proteins constituted the majority of nodes, lipids were consistently integrated within each module rather than forming isolated clusters, highlighting the tight biochemical coupling of the EV multi-ome. Functional annotation of protein components within each module identified distinct biological hallmarks of preterm development (**Fig. 5d**). Module M1, the largest community, reflected a systemic shift in neonatal metabolism through its enrichment in macromolecule catabolism and intracellular organization, effectively capturing the progressive decline of growth-associated cargo as the infant adapts to the extrauterine environment. The maturation of the neonatal immune system was divided into two distinct programs: module M2 was characterized by endocytosis and complement-mediated responses, hallmarks of the innate immune system, while module M3 focused on the expansion of the adaptive immune repertoire, specifically immunoglobulin-mediated signaling. The remaining modules highlighted the biosynthetic and structural demands of development; module M4 was associated with ribosomal biogenesis and cytoplasmic translation, whereas module M5 centered on extracellular matrix organization and tissue remodeling, suggesting a direct link between EV molecular cargo and the maturation of systemic tissue architecture.

### Association of integrated EV molecular modules with clinical phenotypes

To evaluate the clinical relevance of the integrated EV programs, we interrogated the associations between cross-omics molecular modules and neonatal outcomes using a CAMERA-based competitive gene set framework (**Fig. 6a**). By incorporating linear mixed-effects modeling with duplicate correlation, we accounted for the longitudinal nature of the data while adjusting for gestational age at birth. This module-level strategy increased statistical power to detect coordinated molecular programs that remained below the significance threshold in single-feature analyses.

**Fig. 6:**
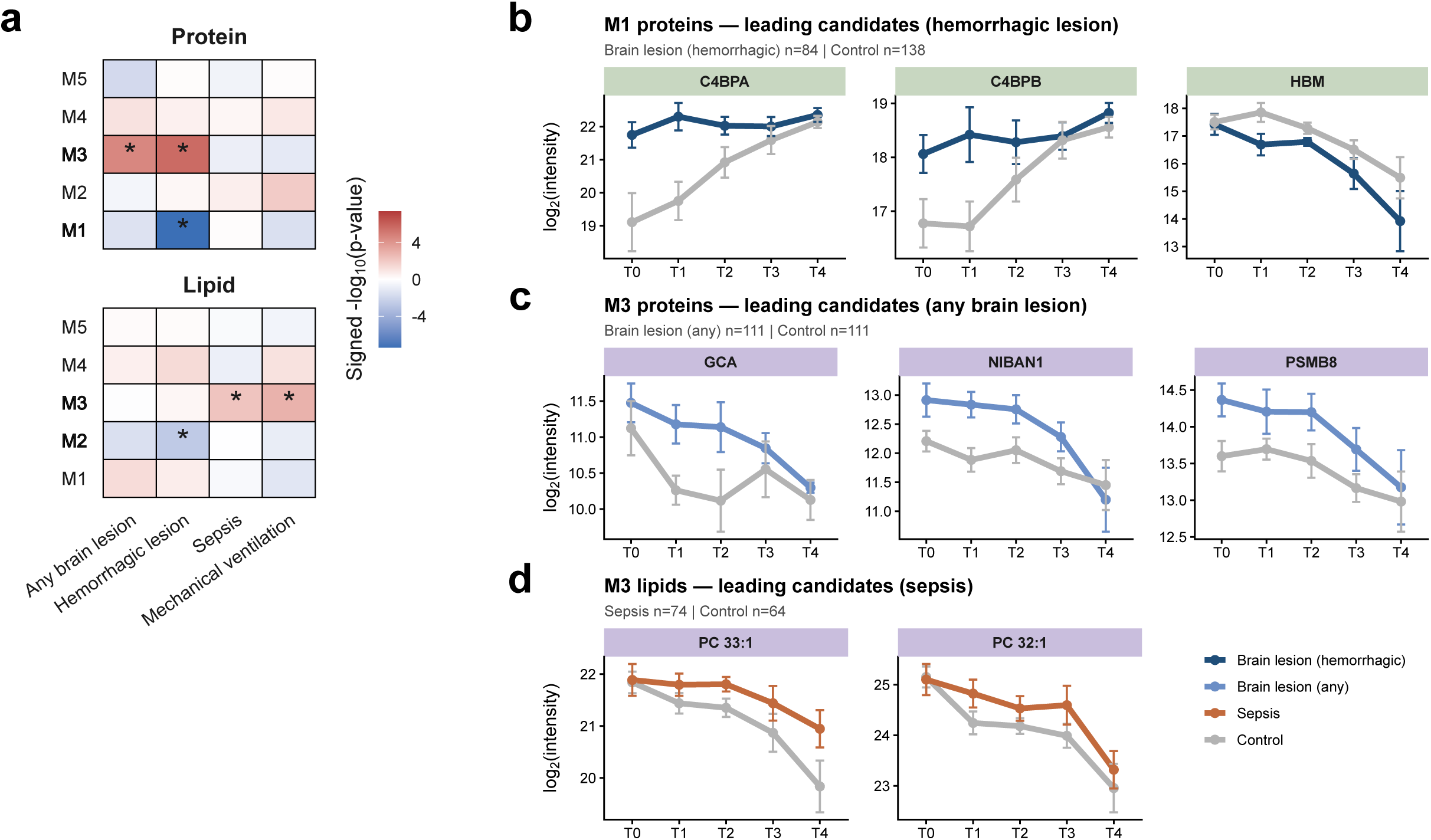
Integrated EV modules show significant associations with clinical phenotypes in preterm infants. (a) Heatmap summarizing module-level associations between EV molecular modules and clinical outcomes, assessed using a CAMERA-based competitive gene set framework. Protein (top) and lipid (bottom) groups within each module were tested separately. Colors represent signed −log10(p-values), with red indicating positive associations and blue negative associations; asterisks denote associations significant after global FDR correction. Any brain lesion includes both hemorrhagic brain injury and white matter injury. While only a limited number of individual lipid species reached statistical significance after multiple testing, coordinated shifts across lipid modules were detected at the module level, highlighting the ability of module-based approaches to capture distributed but consistent EV molecular patterns. (b–d) Longitudinal trajectories of representative leading-edge candidates illustrating the molecular programs underlying significant module–outcome associations. Lines represent mean ± s.e.m. across timepoints (T0–T4).

Module M1 displayed a highly significant association with hemorrhagic brain lesions (FDR = 1.6 × 10⁻□). Although the global module trajectory reflected a progressive decline in metabolic and proteostatic cargo, its leading-edge signal was driven by a compensatory increase in complement regulators C4BPA (β = 1.44) and C4BPB (β = 0.82), contrasted with a marked depletion of the hemoglobin subunit HBM (β = −0.74) (**Fig. 6b**). This internal divergence suggests selective remodeling of EV cargo in the context of hemorrhagic injury, with complement modulation emerging as a dominant adaptive feature.

In parallel, Module M3 exhibited consistent positive associations with both hemorrhagic lesions and the broader category of any brain lesion, including white matter injury (FDR = 3.7 × 10⁻□) (**Fig. 6a,c**). This module reflected a sustained neuroinflammatory and adaptive immune signature, characterized by coordinated upregulation of immunoglobulin families and stress-response mediators such as NIBAN1 (β = 0.55) and the immunoproteasome subunit PSMB8 (β = 0.50), alongside reduced levels of the adhesion molecule SELL (β = −0.49). These trajectories indicate altered leukocyte activation and trafficking dynamics associated with brain injury.

The module-level framework further uncovered concerted lipidomic shifts that were not detectable at the single-feature level (**Fig. 6a,d**). Module M2 lipids were negatively associated with hemorrhagic lesions (FDR = 0.02), reflecting coordinated reductions in membrane-related species, including ether-linked phosphatidylcholines such as PC O-37:8 (β = −0.96) and PC O-39:8 (β = −0.79). Conversely, lipid components within module M3 were positively associated with systemic stress phenotypes, including sepsis (FDR = 0.04) and mechanical ventilation (FDR = 0.01). Representative species such as PC 33:1 and PC 32:1 showed sustained elevations in affected infants (**Fig. 6d**), accompanied by increases in diacylglycerols, consistent with lipid mobilization during inflammatory and respiratory stress.

Collectively, these results demonstrate that clinical phenotypes in very preterm infants are more robustly captured at the level of integrated EV molecular modules than individual features. Notably, the cross-layer convergence of Module M3 across proteins and lipids reinforces a coordinated link between adaptive immune activation and systemic inflammatory outcomes, highlighting the value of multi-omics integration to resolve distributed biological programs underlying neonatal pathology.

## Discussion

Very preterm birth displaces a critical phase of third-trimester maturation into an extrauterine environment characterized by abrupt shifts in oxygen exposure, inflammation, nutrition and intensive care interventions. Longitudinal systems studies have shown that postnatal immune development follows highly ordered and reproducible trajectories, even in preterm infants^6,16,17^. Dissecting intrinsic developmental timing from superimposed clinical stress therefore requires molecular readouts that are both temporally sensitive and biologically specific. Here, we demonstrate that plasma EVs provide such a compartment. By extending Mag-Net enrichment^14^ to enable integrated proteomic and lipidomic profiling from 10 µL plasma, and applying this framework from birth to term-equivalent age, we reconstruct structured EV trajectories and identify cross-omics modules associated with clinically relevant phenotypes, most prominently brain injury.

A first advance is methodological, with direct implications for biological interpretation. Plasma EV profiling is constrained not only by minimal input volumes but also by incomplete separation from abundant non-EV particles, particularly lipoproteins. High-resolution plasma fractionation studies now define conserved EV protein and lipid features alongside non-EV hallmarks^13^, and demonstrate that lipoproteins can physically associate with EVs, forming hybrid nanoparticle complexes^18^. Methodological work further shows that combined size- and density-based approaches improve purity but cannot completely eliminate co-isolates^19^. Within this context, extracting proteins and lipids from the same EV preparation preserves the intrinsic coupling between membrane composition and protein cargo, strengthening cross-layer interpretation while acknowledging known plasma complexities.

Biologically, EV remodeling across early postnatal life is highly structured. More than half of quantified EV proteins display significant temporal dynamics, resolving into two dominant programs: a birth-enriched profile dominated by translation and RNA metabolism, and a progressively increasing immune program enriched for complement and immunoglobulin-associated functions. This transition mirrors systems-level neonatal studies demonstrating rapid, time-driven immune reorganization, with complement pathways among the most reproducibly regulated components in early life^6,20^. In large extremely preterm cohorts, postnatal age explains a substantial proportion of circulating proteomic variance, exceeding gestational age effects beyond the earliest window^17^. The EV trajectories observed here are therefore consistent with a broader temporal architecture of immune ontogeny, but captured within a selectively packaged vesicular compartment rather than bulk plasma.

The immune “ramp” we observe is not generic. Longitudinal plasma proteomics demonstrates coordinated early increases in classical and terminal complement components, rising IgM, and declining maternally derived IgG during the first week of life^20^. Our EV modules enriched for complement and immunoglobulin-linked programs align with these dynamics, suggesting that EV cargo reflects structured maturation of innate and adaptive immune interfaces rather than nonspecific inflammation.

The EV lipidome follows a complementary but distinct pattern. Although fewer lipid species are time-structured, remodeling is clearly species- and structure-dependent. Enrichment toward TEA of triacylglycerols and ether-linked phosphatidylcholines, together with chain-length and linkage-specific redistribution within membrane classes, argues against uniform class-wide drift. Ether lipids are mechanistically linked to exosome biogenesis: modulation of ether-lipid synthesis alters vesicle release and composition^21^, and plasmalogen enrichment influences membrane biophysical properties^22^. These data support the interpretation that ether-PC increases may reflect regulated membrane remodeling during EV maturation. At the same time, neutral lipid signals must be interpreted cautiously. LDL particles can co-purify and associate with EVs^18,23^, and TG species are enriched in non-EV plasma fractions^13^. Conversely, lipid-rich exosome-sized vesicles with immuno-metabolic functions have been described^24^. Our interpretation therefore remains balanced: TG enrichment may reflect both regulated EV-associated biology and the evolving nanoparticle environment of neonatal plasma.

Integration of proteomic and lipidomic layers provides the clearest insight. By restricting network construction to temporally regulated features and modeling cross-layer correlations, we identify five modules summarizing coordinated longitudinal behavior. Network-based dimensionality reduction reduces multiple testing burden and yields biologically coherent units that are more stable than single features in small, heterogeneous cohorts. In our dataset, modules partition immune maturation into distinct programs and capture distributed lipid shifts that are not evident at the single-molecule level. This module-level organization transforms high-dimensional data into interpretable biological states.

Clinically, module-phenotype associations converge on brain injury. Complement provides a biologically plausible bridge between systemic inflammation and immature white matter vulnerability. In neonatal models, activation of the C3a-C3aR axis impairs oligodendrocyte precursor maturation and promotes hypomyelination^25^. Genetic disruption of classical pathway components such as C1q reduces hypoxic-ischemic brain injury in neonatal models^26,27^, while C5a signaling contributes to cortical injury in experimental preterm birth^28^. Complement components also participate in glia-mediated neurotoxicity, including C1q-driven astrocyte activation^29^. These mechanistic data support the plausibility that EV modules enriched for complement components reflect systemic states linked to brain vulnerability.

Translational work further supports EV-mediated inter-organ communication in prematurity. In preterm infants, higher oxygen exposure associates with increased circulating EV cargo containing inflammasome components, and adoptive transfer of such EVs induces lung and brain injury phenotypes in neonatal models^30^. EV cargo has also been linked to bronchopulmonary dysplasia severity with functional validation^31^. Together, these findings support the concept that circulating EVs can report and potentially mediate lung-immune-brain interactions. Within this framework, associations between our EV modules, respiratory support and brain injury are biologically coherent.

An additional interpretive layer concerns the plasma protein corona. EVs incubated in plasma acquire surface-associated proteins, including complement components and immunoglobulins^32^. This is particularly relevant in plasma, where EV surface-associated proteins may represent vesicle biology and the surrounding environment simultaneously. Importantly, such coronas modulate immune cell uptake and trafficking^33^. Complement- and Ig-rich modules may therefore reflect a combination of intrinsic cargo remodeling and altered opsonization dynamics, both relevant to neonatal inflammation and sepsis. Together, these observations indicate that immune-enriched EV modules provide a structured representation of vesicle-associated immune remodeling.

Several limitations define the scope of inference. Plasma EV preparations remain heterogeneous in subtype and cellular origin, and non-EV contributions cannot be fully excluded, particularly in lipid space. Our aim was to characterize an EV-enriched systemic compartment rather than assign tissue specificity, although EV proteomics has demonstrated tissue-enriched signatures in plasma under optimized conditions^34^. Nutritional transitions plausibly influence both immune and lipid trajectories. Although longitudinal mixed-effects modeling and module-level statistics mitigate limited cohort size and unbalanced late sampling, independent validation will be required before clinical translation.

In summary, integrated EV proteomic–lipidomic profiling from 10 µL of plasma captures coherent, time-structured molecular programs in very preterm infants. By enabling parallel recovery of proteins and lipids from the same micro-volume preparation, this workflow preserves cross-layer biological relationships under sampling conditions that are realistic in the NICU setting. EV proteins trace a coordinated shift from growth-associated translation toward immune competence, consistent with established patterns of neonatal immune ontogeny^6,16^. EV lipids undergo selective, structure-dependent remodeling compatible with regulated membrane biology^21^, while remaining interpretable within the known constraints of plasma nanoparticle complexity. Cross-omics modules condense these longitudinal dynamics into stable and biologically meaningful units and reveal clinically relevant programs associated with brain injury and systemic stress. Together, these findings demonstrate that ultra-low-volume EV multi-omics is not only technically feasible, but capable of resolving structured developmental and injury-related trajectories, providing a scalable framework for longitudinal molecular profiling in neonatal medicine.

## Methods

### Cohort description

This prospective, longitudinal study included 16 very preterm infants born at <32 weeks of gestational age and admitted to the Neonatal Pathology and Intensive Care Unit of IRCCS Istituto Giannina Gaslini (Genoa, Italy). Infants were consecutively enrolled between November 2022 and November 2023 and followed from birth to term-equivalent age (40 weeks postmenstrual age) according to a predefined study protocol.

A total of 74 plasma samples were collected across five scheduled timepoints (T0-T4). Only infants with samples available at a minimum of four timepoints were included in longitudinal analyses. Demographic and clinical data were collected from electronic medical records. Baseline characteristics were selected a priori based on relevance to neonatal maturation and availability in the clinical dataset. A summary of demographic and clinical characteristics is provided in **Table 1**. Neurological status was assessed using routine clinical neuroimaging, including cranial ultrasound and, when clinically indicated, magnetic resonance imaging. Brain injury was defined as the presence of any documented brain lesion and recorded as a binary variable. When present, lesions were classified descriptively as hemorrhagic or white matter injury.

The study was conducted in accordance with the Declaration of Helsinki and approved by the Liguria Regional Ethics Committee (approval ID: 469/2021 – DB ID 11730). Written informed consent was obtained from the parents or legal guardians prior to enrollment.

### Nutritional management

All infants were managed according to a standardized institutional nutritional protocol applied consistently across the cohort. Parenteral nutrition was initiated immediately after birth with amino acids and 10% glucose. At approximately 24 hours of life, a multicomponent lipid emulsion (SMOFlipid; soybean oil, medium-chain triglycerides, olive oil, and fish oil) was introduced. Minimal enteral feeding with maternal milk, or pasteurized donor milk when maternal milk was unavailable, was initiated within the first 24 hours of life. Upon reaching 80 mL/kg/day of enteral intake, human milk was fortified (Aptamil BMF) to provide additional protein and long-chain polyunsaturated fatty acids, including docosahexaenoic acid (DHA) and arachidonic acid (AA). Parenteral lipid administration was discontinued when enteral intake reached 90-100 mL/kg/day. Amino acids and glucose infusion were adjusted according to standard clinical practice.

### Plasma sample collection

Venous or capillary blood samples (0.5-1 mL) were collected longitudinally at five predefined timepoints and integrated with routine clinical care to minimize procedural burden. The first sample (T0) was obtained within 40 minutes after birth during placement of the umbilical venous catheter. Subsequent samples were collected at 48-72 hours of life (T1), 7 days of life (±1 day; T2), approximately 33 weeks postmenstrual age (T3), and at term-equivalent age (∼40 weeks postmenstrual age; T4). When available, the umbilical venous catheter was used for early sampling; otherwise, peripheral venipuncture was performed. Samples at T3 and T4 were obtained via peripheral venipuncture.

Whole blood was collected into EDTA tubes and processed within 3 hours of collection. Plasma was isolated by centrifugation at 1200 × g for 15 minutes at room temperature. The supernatant was aliquoted, anonymized using alphanumeric identifiers, and stored at −80 °C until extracellular vesicle enrichment. Samples underwent a single freeze-thaw cycle for analysis.

For method optimization and technical validation experiments, anonymized pooled plasma from healthy adult donors was used. Individual plasma units were combined to generate homogeneous pooled material, which was aliquoted and processed in four independent technical replicates under identical experimental conditions. These pooled samples were used exclusively for workflow benchmarking and reproducibility assessment and were not included in longitudinal clinical analyses.

### Automated extracellular vesicle enrichment and integrated proteomic-lipidomic sample preparation

EVs were enriched from plasma using a semi-automated SAX magnetic bead workflow adapted from the Mag-Net method^14^ to enable parallel proteomic and lipidomic extraction from minimal input volumes. All steps were performed on a KingFisher™ Apex magnetic handling station (Thermo Fisher Scientific) at 4 °C.

For each sample, 10 µL of plasma were thawed on ice and mixed with 40 µL PBS and 50 µL Bis-Tris Propane (BTP) binding buffer (100 mM BTP, pH 6.3, 150 mM NaCl), yielding a final volume of 100 µL. EV capture was performed using 10 µL of MagReSyn™ SAX magnetic beads (ReSyn Biosciences), pre-equilibrated in BTP wash buffer (50 mM BTP, pH 6.5, 150 mM NaCl). Sequential washes were performed to remove non-vesicular plasma components.

Bead-bound EVs were subjected to monophasic extraction using 600 µL 1-butanol/acetonitrile/water (3:1:1, v/v)^15^. This step promotes membrane disruption and the subsequent release of the molecular cargo. Samples were incubated overnight at −20 °C to induce protein precipitation and aggregation capture (PAC), while lipid species remained in the soluble organic phase.

The following day, samples were reprocessed on the KingFisher™ Apex platform. During the initial protein aggregation capture (PAC) step, the precipitated proteins were retained on the magnetic beads, while the lipid-rich supernatant was recovered and processed separately. The lipid fraction was clarified by centrifugation (4 °C, 4,000 × g, 25 min), transferred to clean tubes, vacuum-dried, and stored at −20 °C until LC-MS/MS analysis.

In parallel, the bead-bound proteins underwent automated processing, including sequential washes with acetonitrile (ACN), 70% ethanol, and isopropanol (IPA) to remove contaminants. On-bead digestion was performed in 100 mM Tris-HCl (pH 8.0) containing 10 mM tris(2-carboxyethyl)phosphine (TCEP) and 4 mM chloroacetamide (CAA) for reduction and alkylation. Proteins were digested at 37 °C for 4 h using LysC (Wako) and trypsin (Promega) at enzyme-to-protein ratios of 1:400 and 1:200 (w/w), respectively. Peptides were acidified with 10 µL of 2% (v/v) trifluoroacetic acid (TFA) and stored at −20 °C until LC-MS/MS analysis.

### Data acquisition

#### Lipidomic LC-MS/MS acquisition

Lipidomic analyses were performed using a Vanquish Horizon UHPLC system coupled to an Orbitrap Fusion™ Tribrid™ mass spectrometer (Thermo Fisher Scientific). Dried lipid extracts were reconstituted in 1-butanol (BuOH)/isopropanol (IPA)/water (H₂O) (8:23:69, v/v) supplemented with 5 mM phosphoric acid, and 5 µL of each sample were injected onto a reversed-phase column.

Chromatographic separation was achieved on an ACQUITY UPLC BEH C18 column (2.1 × 50 mm, 1.7 µm; Waters) maintained at 65 °C, using a 7-minute gradient at a flow rate of 600 µL/min. Mobile phase A consisted of 10 mM ammonium formate in 60:40 ACN/H_2_O with 0.1% formic acid (FA); mobile phase B consisted of 10 mM ammonium formate in 90:10 IPA/ACN with 0.1% FA. The gradient started at 15% B, increased to 30% B over 1 minute, then to 100% B over the following 5 minutes, and was held at 100% B for 1 minute. The column was subsequently re-equilibrated for 1 minute.

Eluting lipid species were analyzed by electrospray ionization tandem mass spectrometry (ESI-MS/MS). For technical validation experiments, data were acquired in both positive and negative ionization modes using a spray voltage of ±3.5 kV. For the longitudinal clinical dataset, acquisition was performed in positive ion mode only (+3.5 kV). The inlet capillary temperature was set to 300 °C. Nitrogen was used as sheath and auxiliary gas at flow rates of 30 and 10 arbitrary units (AU), respectively.

Data were acquired in data-dependent acquisition (DDA) mode, alternating full MS and MS/MS scans. Full MS survey scans were acquired in the Orbitrap analyzer over a mass range of 70-1000 m/z at 70,000 resolution (at 200 m/z), with an AGC target of 1 × 10□ and a maximum injection time of 100 ms. Up to five MS/MS events were triggered per MS scan using an isolation window of 1.4 Da and an intensity threshold of 1.6 × 10□. Fragment ion spectra were acquired at 17,500 resolution, with an AGC target of 1 × 10□ and a maximum injection time of 50 ms. Previously fragmented precursor ions were dynamically excluded for 2 seconds. Higher-energy collisional dissociation (HCD) was performed using stepped normalized collision energies of 20, 30, and 40.

#### Proteomic LC-MS/MS acquisition

Proteomic analysis was performed using an Evosep One chromatographic system coupled to an Orbitrap Exploris™ 480 mass spectrometer (Thermo Fisher Scientific) equipped with a FAIMS Pro Duo interface.

Peptide digests (20 µL) were loaded onto EvoTips according to the manufacturer’s instructions. Peptide separation was achieved on an Aurora Rapid column (8 cm x 75 μm; IonOpticks) maintained at 25 °C, using the pre-programmed 60 samples per day (60 SPD) method at a constant flow rate of 1 µL/min.

Data were acquired in data-independent acquisition (DIA) mode. Full MS scans were acquired at 60,000 resolution over 380-980 m/z, with an automatic gain control (AGC) target of 300% and the maximum injection time (IT) set to Auto. MS/MS acquisition consisted of 50 DIA windows (15 Da width with 1 Da overlap) acquired at 15,000 resolution, with injection time set to Auto and a normalized collision energy of 25%. The FAIMS compensation voltage was set to −45 V at standard resolution. Data were acquired in profile mode using positive ion polarity.

### Data processing and quality control

#### Lipidomic data processing

Raw lipidomics data were processed using MS-DIAL v5.5 for peak detection, deconvolution, retention time alignment, and lipid annotation^35^. The resulting master area table was refined using MS-FLO v1.8 to perform additional deisotoping and to resolve redundant or duplicated lipid features, thereby improving annotation consistency^36^.

The curated table was imported into R v4.5.2 for downstream processing. Putative contaminants (e.g., chemical reagents, plasticizers, polymer-related compounds, and background signals) were excluded based on annotation keywords and pattern-based filtering.

Technical reproducibility was assessed using pooled quality control (QC) samples. For each lipid feature, mean intensity and coefficient of variation (CV) were calculated across QC injections. Features with CV >30% were removed.

To retain biologically informative species in the longitudinal dataset, a fill-rate filter was applied. Lipid features were retained if they exhibited ≥70% non-missing values in at least one timepoint. When multiple features corresponded to the same lipid annotation, a single representative feature was retained based on highest fill rate, lowest QC CV, and highest mean QC intensity.

Intensity values were log₂-transformed and median-centered to correct for sample-wise technical variability. Missing values were imputed assuming left-censored missingness using a Gaussian distribution downshifted by 1.8 standard deviations with a width of 0.3 standard deviations.

#### Proteomic data processing

Proteomic data processing was performed separately for technical validation experiments and for the longitudinal clinical dataset using Spectronaut v18 (Biognosys AG)^37^.

For technical validation experiments, raw data were analyzed using the Method Evaluation workflow to assess identification depth and analytical reproducibility.

For the longitudinal clinical dataset, raw files were processed in directDIA mode with default settings. Searches were performed against the UniProt human reference proteome with Trypsin/P and LysC specified as cleavage enzymes, carbamidomethylation (C) as a fixed modification, and methionine oxidation and protein N-terminal acetylation as variable modifications. False discovery rate was controlled at 1% at peptide spectrum match (PSM), peptide, and protein group levels using a target–decoy strategy. Protein inference and filtering were performed using the Identified (Q-value) setting, and protein quantification was based on MS2-level intensities.

Only the longitudinal dataset underwent downstream data curation in R (v4.5.2). Protein groups were harmonized to a single representative UniProt accession and gene symbol, prioritizing reviewed Swiss-Prot entries when available. Proteins were retained if they exhibited ≥70% non-missing values in at least one timepoint. Intensities (log₂ scale) were median-centered across samples to correct for sample-wise technical variability.

Missing values in the longitudinal dataset were handled using a hybrid imputation strategy reflecting different plausible missingness mechanisms in MS-based proteomics.

As missing-at-random (MAR) and missing-not-at-random (MNAR) mechanisms cannot be directly identified from observed data alone, MNAR was operationally assumed when missingness patterns were consistent with left-censoring. Specifically, within each timepoint block, proteins exhibiting very low detection rates were classified as MNAR (proportion of observed values <0.30 when ≥12 samples were available in that timepoint, or <0.40 when <12 samples were available). These thresholds were selected to conservatively identify features with systematic under-detection while avoiding over-classification of sparse but potentially informative signals. Missing values not flagged as MNAR were treated as MAR. MAR values were imputed using structured least squares adaptive (SLSA) imputation (*imp4p* R package), whereas MNAR values were imputed from a downshifted Gaussian distribution (mean −1.8×SD, SD×0.3) to model signal intensities below the limit of detection.

### Data analysis

All statistical analyses were performed in R v4.5.2. Proteomic and lipidomic datasets were analyzed separately and subsequently integrated using a network-based multi-omics framework. Longitudinal molecular changes were assessed using per-feature linear mixed-effects models fitted with the *lmerTest* package. For each protein or lipid, abundance was modeled as a function of sampling timepoint (T0-T4) and gestational age at birth, with infant identity included as a random intercept to account for repeated measurements: abundance ∼ timepoint + ga_birth + (1 | infant_id). Timepoint was treated as a categorical factor with T0 as the reference level. Models were fitted using maximum likelihood (REML = FALSE; optimizer = bobyqa).

A global time effect was assessed using Type-III ANOVA on the timepoint term, as implemented in *lmerTest*. For endpoint analyses, the contrast between term-equivalent age (T4) and birth (T0) was extracted from the model as the coefficient for timepoint T4. Effect sizes were reported as β coefficients, representing model-estimated differences in log₂-transformed abundance. For timepoint effects, β corresponds to the log₂ fold-change relative to the reference timepoint (T0), adjusted for gestational age at birth. P-values were adjusted using the Benjamini-Hochberg procedure. Features with FDR_global < 0.05 were considered significantly time-regulated. Features were considered significantly regulated at T4 versus T0 if they met FDR_T4vsT0 < 0.05 and |β| > 0.58, corresponding to an approximate 1.5-fold change on the original abundance scale, thereby prioritizing biologically meaningful shifts.

Cross-omics integration was performed using temporally regulated features (FDR_global < 0.05). Protein and lipid abundance profiles were z-score standardized, and pairwise Spearman correlations were computed across matched longitudinal samples. Correlation p-values were corrected using BH adjustment. Network edges were defined by FDR < 0.05 and |ρ| ≥ 0.35. The resulting weighted, undirected network was partitioned using Louvain community detection to identify cross-omics modules. Modules with fewer than 10 total features were excluded from downstream analyses.

Associations between molecular modules and clinical outcomes were tested separately for proteins and lipids using CAMERA gene-set testing within the *limma* framework. Models included the clinical variable of interest as the main effect, with timepoint and gestational age at birth as covariates. Repeated measurements were accounted for using duplicateCorrelation with infant identity as the blocking factor. P-values were corrected for multiple testing across all module-outcome-layer combinations using the BH procedure. Associations with FDR < 0.05 were considered statistically significant.

Functional enrichment of proteomic clusters and network modules was performed in R using Gene Ontology Biological Process and Reactome pathway databases, using all quantified proteins as background and Benjamini–Hochberg correction (FDR < 0.05). Lipid ontology enrichment was performed using the LION web-based tool, ranking lipid species according to their T4 versus T0 β coefficients and retaining significantly enriched terms (FDR < 0.05).

## Data Availability

The mass spectrometry proteomics and lipidomics data generated in this study have been deposited to the ProteomeXchange Consortium via the PRIDE partner repository.The datasets will be released publicly once the work is accepted by a peer-reviewed journal.

https://www.ebi.ac.uk/pride/

## Data availability

The mass spectrometry proteomics data have been deposited to the ProteomeXchange Consortium via the PRIDE partner repository^38^ with the dataset identifier PXD074778. *During peer review, the dataset can be accessed using the following credentials: Username; Password: Ap7CxD6BMgE6; or via the access token: mMjU7VHYzPW2. These reviewer credentials will be removed prior to publication*.

## Code availability

The custom R scripts used to generate the results reported in this study are available from the corresponding author upon request.

## Author contributions

C.L. and A.P. conceived and designed the study. N.G. and F.A. performed the experiments and curated the dataset. N.G. optimized the workflow, conducted the bioinformatic and statistical analyses, and drafted the manuscript. F.A. and M.Ba. contributed to data interpretation. L.R. and S.S. contributed to sample processing and data acquisition. G.T., C.A., F.V., and L.A.R. contributed to patient recruitment, clinical data collection, and clinical interpretation. G.L. and M.Bru. contributed to data interpretation and critically revised the manuscript. L.A.R., C.L., and A.P. jointly supervised the study. All authors reviewed and approved the manuscript.

## Competing interests

The authors declare no competing interests.

## Notes

### Competing Interest Statement

The authors have declared no competing interest.

### Funding Statement

Ministry of Health (5M-2018-23680427, to A.Pe.)

### Author Declarations

The study was conducted in accordance with the Declaration of Helsinki and was approved by the Liguria Regional Ethics Committee (approval ID: 469/2021 DB ID 11730).

### Summary of Updates

Updated author list and affiliations; additional analyses and minor revisions to the manuscript text and figures.

