## Supplementary figures for "Small volumes, deep insights: longitudinal plasma EV multi-omics in very preterm infants"

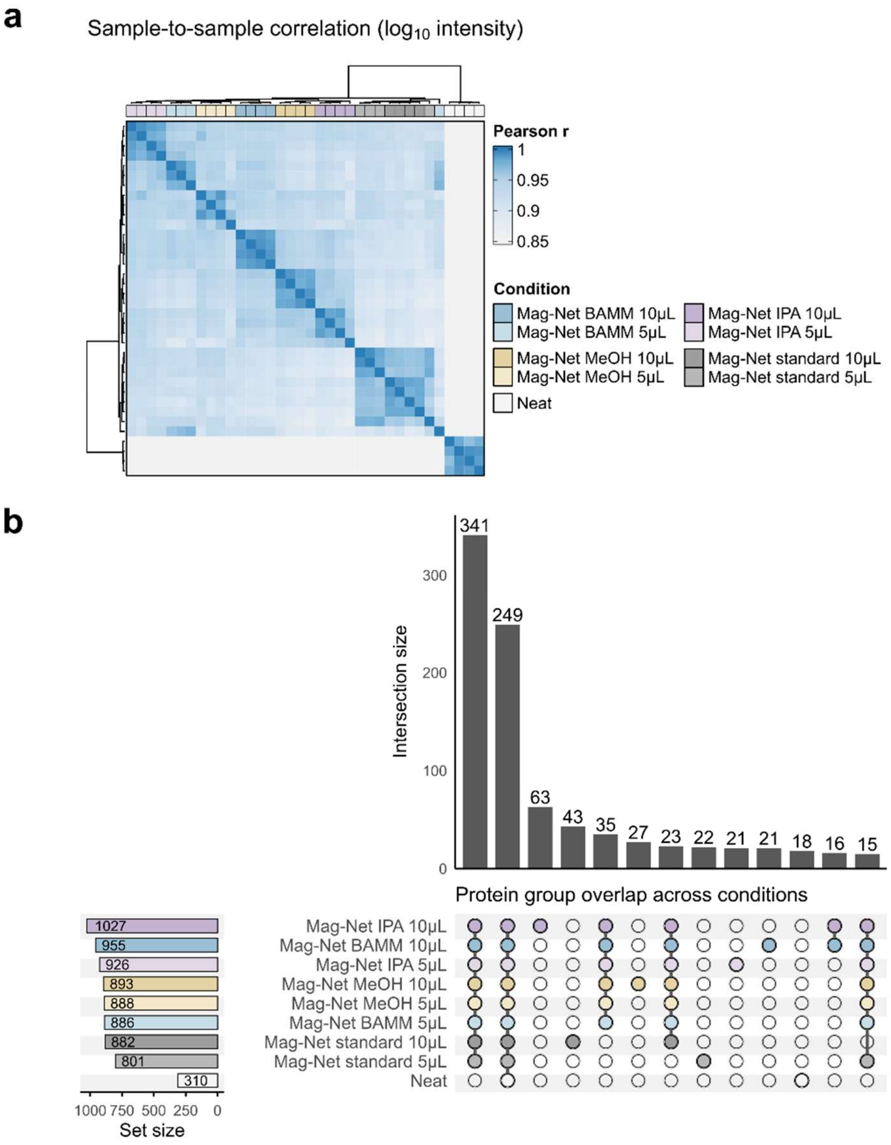


**Supplementary Fig.S1: Reproducibility and proteome overlap across EV enrichment and extraction conditions.** (a) Sample-to-sample Pearson correlation matrix based on $log_{10}$-transformed protein intensities across all technical conditions, including Mag-Net standard, BAMM, isopropanol (IPA), methanol (MeOH), and direct plasma digestion (“Neat”), at 5 µL and 10 µL inputs. Hierarchical clustering demonstrates high concordance among EV-enriched preparations, with clear separation from direct plasma digestion. (b) Protein group overlap across extraction strategies and plasma input volumes. The UpSet plot summarizes shared and unique protein identifications among conditions. Bars (top) indicate the size of each intersection; horizontal bars (left) show total protein group counts per condition. The majority of identified proteins are consistently detected across EV-enriched workflows, supporting preservation of the core EV-associated proteomic landscape under micro-volume conditions.

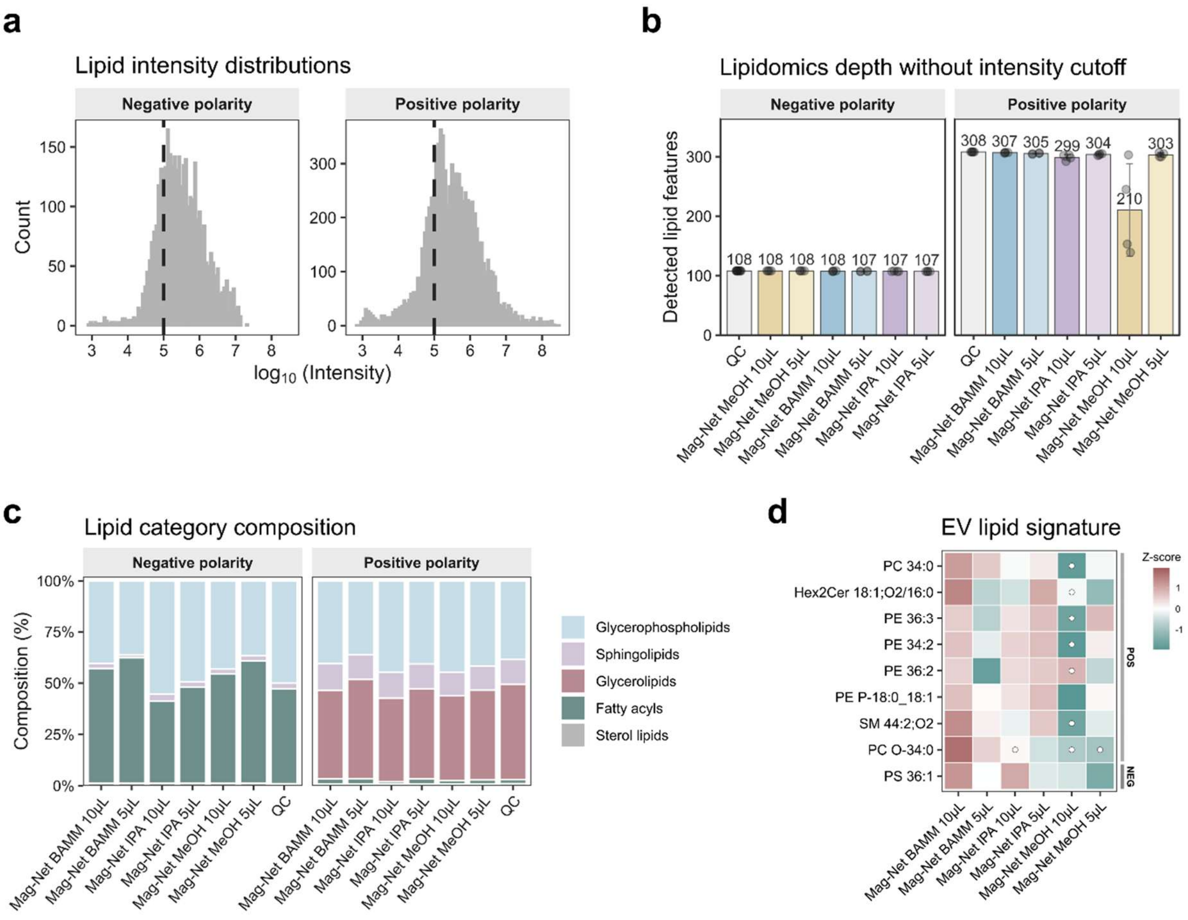


**Supplementary Fig. S2: Lipidomic performance and compositional consistency across EV extraction strategies.** (a) Distribution of $\log_{10}$-transformed lipid intensities in negative and positive ionization modes across EV-enriched preparations. The dashed vertical line indicates the intensity threshold ($10^5$) applied for comparative evaluation of high-confidence lipid features. (b) Total number of detected lipid species in negative and positive polarity without application of an intensity threshold. Bars represent individual technical replicates; numbers above bars indicate total detected features. Overall lipidomic depth is largely comparable across extraction strategies and plasma input volumes. (c) Relative lipid class composition across conditions in negative and positive polarity. Stacked bar plots show the proportional distribution of major lipid categories, demonstrating stable class-level composition independent of solvent choice or plasma input. (d) Heatmap of selected conserved EV-associated lipid features from the density-gradient reference atlas (Rai et al.). Values are row-wise z-scores of log-transformed intensities across conditions. Conserved EV lipid features are consistently detected across EV-enriched workflows.

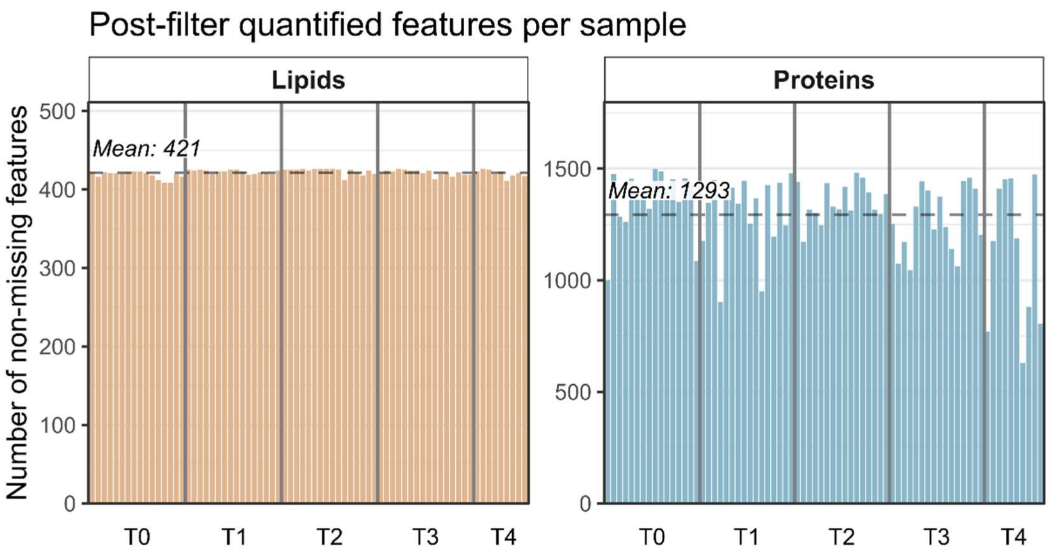


**Supplementary Fig. S3: Post-filter quantification depth per sample across the longitudinal cohort.** Bars represent the number of non-missing features per sample after application of the timepoint-aware fill-rate filter (≥70% observed values in at least one timepoint), applied consistently to both proteomic and lipidomic datasets prior to downstream analyses. Samples are ordered by timepoint (T0-T4). Dashed lines indicate the mean number of quantified features per layer. Lipidomic depth remained highly stable across timepoints (mean = 421 features per sample), whereas proteomic depth showed greater inter-individual variability (mean = 1,293 features per sample).
